# Monte Carlo Committee Simulation with Large Language Models for Predicting Drug Reimbursement Recommendations and Conditions: A Novel Neurosymbolic AI Approach

**DOI:** 10.64898/2026.03.02.26347434

**Authors:** Ghayath Janoudi, Mara Rada, Eugene Yasinov, Trevor Richter

## Abstract

**Background:** Health technology assessment (HTA) agencies issue reimbursement recommendations that determine patient access to new therapies. Predicting these outcomes would enable sponsors to optimize market access strategies and health systems to anticipate budget impacts. However, traditional machine learning approaches require extensive manual feature extraction and predict only categorical outcomes, not the specific conditions attached to recommendations.

**Methods:** We developed Monte Carlo Committee Simulation, a neurosymbolic system that simulates multi-panelist deliberation using 14 persona-conditioned large language model panelists with weighted voting and uncertainty quantification. We conducted a temporal external validation study on CDA-AMC (Canada’s Drug Agency) sponsor-submitted recommendations published between October 2024 and December 2025 (n=67), after the knowledge cutoff of the underlying models, ensuring predictions reflected reasoning rather than memorization. The system predicted both recommendation category (Reimburse with Conditions, Do Not Reimburse) and five condition categories (Population Restrictions, Prescriber/Setting Requirements, Continuation Conditions, Economic Conditions, Evidence Conditions).

**Results:** On submissions where the system expressed confidence (n=44), recommendation prediction achieved 93.2% accuracy (95% CI: 84.1–100.0%), exceeding the 91.8% (95% CI: 83.7–98.0%) majority class baseline. The system demonstrated superior discrimination versus chance level (AUROC 0.817, 95% CI: 0.45–0.99, vs 0.500) and calibrated confidence estimates (ECE = 0.091). Pre-specified Strength of Mandate stratified accuracy from 96.8% (High, 95% CI: 90.3–100.0%) to 40.0% (Weak, 95% CI: 0.0–80.0%), with 83.3% of errors occurring in cases flagged as uncertain (p=0.0025). Analysis of the 5 abstained cases confirmed 40.0% accuracy, validating the system’s identification of uncertain predictions. For condition prediction, the system achieved 48.8% subset accuracy, requiring correct simultaneous prediction of all 5 condition categories (2^5^ = 32 possible combinations), and 86.3% Hamming accuracy versus 25.8% for a no-conditions baseline. Per-category accuracy ranged from 68.3% (Continuation Conditions) to 97.6% (Economic Conditions), with Continuation Conditions demonstrating the strongest discriminative ability (AUROC 0.896, 95% CI: 0.79–0.98).

**Conclusions:** Monte Carlo Committee Simulation enables a shift from reactive to proactive market access: anticipating specific reimbursement conditions before committee review, with calibrated confidence that identifies which predictions to trust. Validated on temporally separated data the models could not have memorized, the system can be positioned as a forecasting aid that complements rather than replaces human deliberation.

## Introduction

Health technology assessment (HTA) agencies worldwide face the consequential task of evaluating new therapies and making reimbursement recommendations that affect millions of patients. These deliberative processes integrate complex clinical evidence, health economic analyses, patient perspectives, and ethical considerations into decisions that traditionally rely on expert committee judgment.^1,2^ The ability to predict HTA outcomes before formal review would provide significant value for pharmaceutical sponsors planning market access strategies, payers anticipating budget impacts, and health systems preparing for new treatment options. However, developing predictive models for HTA recommendations presents fundamental challenges that distinguish this domain from conventional machine learning applications.

Traditional machine learning approaches struggle with HTA prediction for several interrelated reasons. First, the available datasets are inherently small—major HTA agencies typically issue only dozens to low hundreds of recommendations annually, far below the thousands of examples typically required for robust supervised learning.^3,4^ Second, HTA processes are not static targets; agencies regularly update their deliberative frameworks, adopt new methodological approaches for evidence synthesis, and shift policy priorities in response to evolving healthcare landscapes.^5^ Third, the input features for HTA predictions (e.g. clinical trial results, cost-effectiveness analyses, patient input submissions) exist as lengthy, unstructured documents that resist the standardized feature engineering required by traditional approaches. Recent work has applied machine learning to predict HTA outcomes using extracted features from submission documents, achieving moderate success for binary recommendation prediction.^6^ However, these approaches require extensive manual feature engineering and cannot capture the full evidentiary context that committees consider.

Large language models (LLMs) offer a potential paradigm shift by leveraging their capacity for reasoning over unstructured text and their encoded knowledge of medical, economic, and policy domains.^7,8^ Unlike traditional machine learning, LLMs can process the raw evidentiary documents that HTA committees review (e.g. clinical study reports, pharmacoeconomic analyses, patient group submissions) without extensive feature engineering. Moreover, LLMs present an opportunity to predict not merely categorical reimbursement outcomes, but also the specific conditions attached to recommendations. When HTA agencies recommend reimbursement with conditions (the most common outcome for novel therapies) they specify population restrictions, prescriber requirements, continuation criteria, price reductions, or evidence collection mandates. These conditions constitute the actionable policy output that stakeholders need for implementation planning. While existing literature has developed taxonomies of managed entry agreements and reimbursement conditions retrospectively,^9,10^ no study has attempted prospective prediction of condition types. This represents a critical gap: manufacturers benefit more from knowing “you will likely receive conditional reimbursement with prior therapy requirements and price reduction” than merely “you will likely receive conditional reimbursement.” HTA agencies equally benefit from anticipating which submissions are likely to require managed access arrangements (such as population restrictions, continuation criteria, or evidence collection mandates) given the significant in-house and health system resource implications of administering these conditions.

A critical limitation of applying LLMs directly to prediction tasks is the difficulty of quantifying uncertainty. When an LLM produces a single recommendation prediction, there is no principled way to assess whether the model is highly confident or essentially guessing.^11,12^ This epistemic opacity is particularly problematic in high-stakes domains like HTA, where stakeholders need calibrated probability estimates to inform resource allocation decisions. Furthermore, the deliberative nature of committee decision-making introduces irreducible variability; different committee compositions, meeting dynamics, and framing of evidence can lead to different outcomes even for identical submissions. A single-prompt approach cannot capture this fundamental uncertainty inherent in human deliberation.

By instantiating multiple LLM-based panelists with distinct personas and aggregating their predictions through formal voting rules, we can generate prediction distributions that meaningfully quantify uncertainty. This approach constitutes a *neurosymbolic* architecture: neural components (persona-conditioned LLM panelists) perform evidence interpretation, while symbolic components (weighted voting, convergence criteria, abstention thresholds) perform collective inference with principled statistical properties.^13,14^ The separation allows neural reasoning to handle ambiguity and implicit domain knowledge while symbolic aggregation provides transparency, calibrated uncertainty, and interpretable confidence metrics.

Real HTA committees comprise individuals with diverse expertise (e.g. clinicians, health economists, patient advocates, policy experts) who bring different analytical lenses to the same evidence. This diversity is not noise to be averaged away but rather a feature that enables robust deliberation. Ensemble learning theory demonstrates that combining diverse learners yields predictions that are more accurate and better calibrated than any individual model.^15,16^ By instantiating multiple LLM-based panelists with distinct personas and aggregating their predictions through formal voting rules, we can generate prediction distributions that meaningfully quantify uncertainty.

This design constitutes a neurosymbolic architecture.^13,14^ The neural components (persona-conditioned LLM panelists) interpret unstructured evidentiary documents, drawing on the models’ encoded clinical, economic, and policy knowledge. The symbolic components (voting rules, convergence criteria, and abstention thresholds) aggregate these individual judgments into calibrated probability distributions with principled statistical properties. Neither component alone would suffice: a single LLM prompt cannot quantify its own uncertainty, and purely rule-based systems cannot reason over the lengthy unstructured documents that characterize HTA submissions. The separation keeps reasoning flexible while making aggregation transparent and auditable.

The separation allows neural reasoning to handle ambiguity and implicit domain knowledge while symbolic aggregation provides transparency, calibrated uncertainty, and interpretable confidence metrics.

We propose Monte Carlo Committee Simulation, a neurosymbolic framework that simulates multi-panelist deliberation to predict HTA recommendations and conditions. Our approach samples predictions from 14 persona-conditioned panelists across multiple rounds, generating probability distributions over recommendation outcomes (Reimburse, Reimburse with Conditions, Do Not Reimburse) and—for conditional recommendations—over five condition categories. This two-level prediction framework addresses both what decision will be made and what form that decision will take. The Monte Carlo simulation provides calibrated confidence intervals, enables selective prediction through uncertainty-based abstention, and captures the inherent stochasticity of committee deliberation through a novel “Strength of Mandate” metric that quantifies both inter-round stability and intra-round contestation.

Critically, we conduct all evaluations prospectively on recommendations published after the training data cutoff of our underlying models. This design is essential because LLMs trained on web-scale corpora may have encountered historical HTA recommendations, making it impossible to distinguish between genuine reasoning and memorized recall.^17,18^ By restricting our evaluation to Canada’s Drug Agency (CDA-AMC) sponsor-submitted recommendations published between October 2024 and December 2025, we ensure that our system must reason from evidentiary documents rather than retrieve cached outcomes.

To our knowledge, this represents the first prospective evaluation of LLM-based HTA prediction, the first application of neurosymbolic committee simulation to health technology assessment, and the first attempt to predict the specific conditions attached to reimbursement recommendations.

## Methods

### Study Design

This is a temporal external validation study evaluating predictions of CDA-AMC reimbursement recommendations for sponsor-submitted drug reviews published after the knowledge cutoff of the underlying models. We developed and evaluated a Monte Carlo Committee Simulation system that simulates multi-panelist deliberation to generate probability distributions over recommendation outcomes (Reimburse, Reimburse with Conditions, Do Not Reimburse) and condition categories.

### Data Sources and Eligibility

We included all sponsor-submitted recommendations issued by CDA-AMC’s expert committees (CDEC for non-oncology, pERC for oncology) between October 2024 and December 2025. FMEC (Formulary Management Expert Committee) non-sponsored reviews were excluded due to different document structure and deliberation processes.

Eligibility criteria: - Sponsor-submitted recommendations only (standard HTA pathway) - Post-cutoff recommendations (issued after September 30, 2024)

We split the dataset chronologically: calibration set (October-December 2024) for threshold tuning, and test set (January-December 2025) for final evaluation.

### Document Processing

For each recommendation, we obtained the publicly available executive summary documents from CDA-AMC. Documents were OCR’d from PDF to markdown format. We extracted structured sections (clinical evidence, economic evidence, patient input, committee deliberation) for input to the simulation system.

### Ground Truth

#### Recommendations

Ground truth labels (R/RWC/DNR) were obtained directly from CDA-AMC structured data exports. Time-limited recommendations were classified as RWC (containing evidence collection conditions).

#### Conditions

We developed a 5-category taxonomy with 11 evaluated subcategories for condition classification:

Condition annotations were extracted from “Table 1: Reimbursement Conditions and Reasons” in each document. Primary annotation was performed by GJ; a second reviewer (MR) independently validated 20% of annotations. Discrepancies were resolved through discussion.

**Table 1:** Conditions Categories.

| Category | Subcategories |
| --- | --- |
| Population Restrictions | Biomarker requirement, Line of therapy, Prior treatment requirement, Disease characteristics |
| Prescriber/Setting | Specialist initiation, Treatment centre requirement |
| Continuation Conditions | Maximum treatment duration, Response criteria for continuation |
| Economic Conditions | Comparator-based cost condition, Price reduction required |
| Evidence Conditions | Time-limited approval (real-world evidence collection) |

#### Models

We used two Azure OpenAI models in a mixed-model configuration:

**Table 2:** Large Language Models Specifics.

| Model | Knowledge Cutoff | Version | Role |
| --- | --- | --- | --- |
| GPT-5 | September 30, 2024 | 2025-08-07 | Instance 1 panelists (CDA-AMC Deliberative Framework prompts) |
| GPT-5-mini | May 31, 2024 | 2025-08-07 | Instance 2 panelists (simplified prompts) |

Both models’ knowledge cutoffs predate all recommendations in our evaluation period (October 2024–December 2025), ensuring complete temporal separation between training and evaluation data.

GPT-5 was selected for Instance 1 panelists because the structured CDA-AMC Deliberative Framework prompts require complex multi-domain reasoning across clinical, economic, and policy evidence. GPT-5-mini was used for Instance 2 panelists, whose simplified prompts require less reasoning depth. Using two distinct models also introduces model diversity into the ensemble, reducing the risk of correlated errors across panelists that would arise from a single-model configuration. Both models were chosen to allow sufficient window for collecting temporal validation data pose training cutoff while also allowing the use of more recent and capable models.

All API calls used reasoning at default (medium) to enable stochastic sampling across Monte Carlo rounds.

### Monte Carlo Committee Simulation

#### Committee Composition

Each simulated committee comprises 14 panelists: 7 persona types with 2 instances each.

**Table 3:**
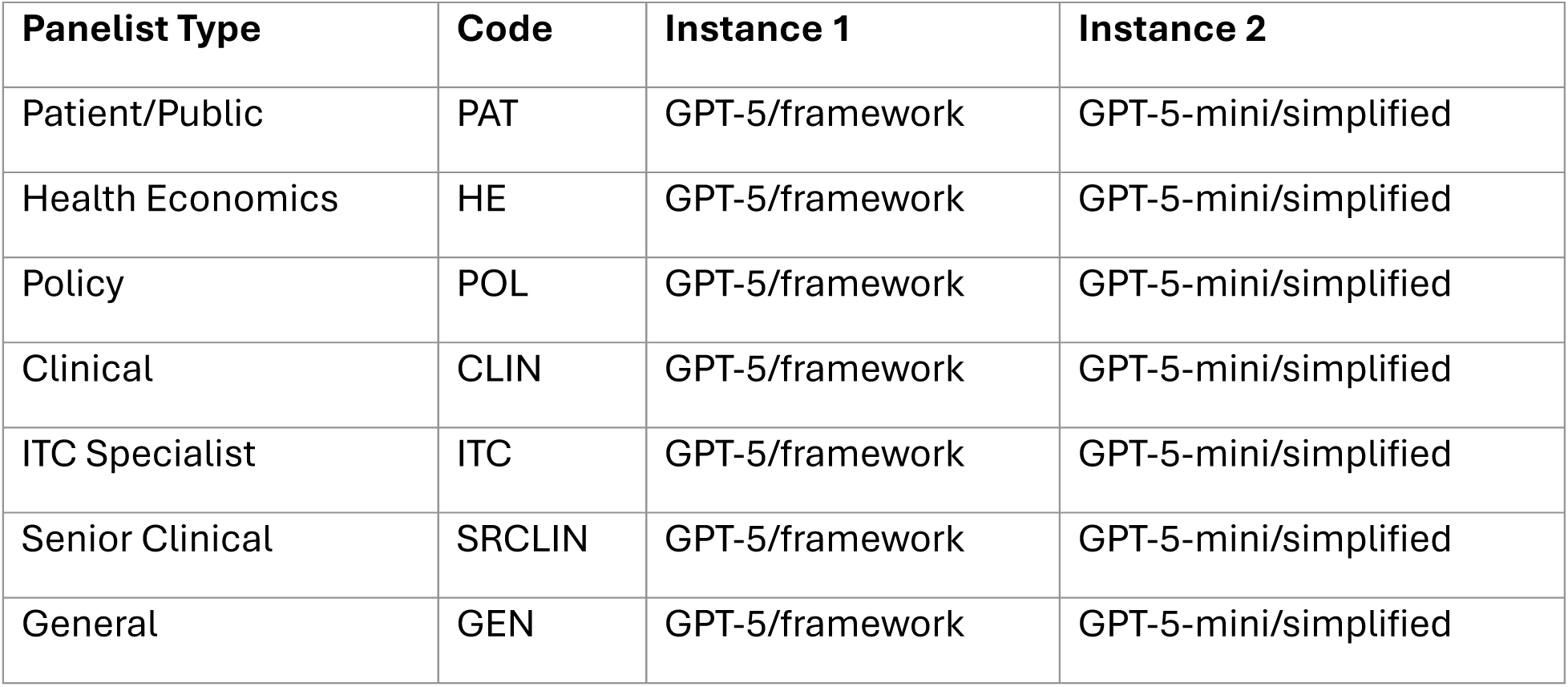
LLM-panelists specifics.

| Panelist Type | Code | Instance 1 | Instance 2 |
| --- | --- | --- | --- |
| Patient/Public | PAT | GPT-5/framework | GPT-5-mini/simplified |
| Health Economics | HE | GPT-5/framework | GPT-5-mini/simplified |
| Policy | POL | GPT-5/framework | GPT-5-mini/simplified |
| Clinical | CLIN | GPT-5/framework | GPT-5-mini/simplified |
| ITC Specialist | ITC | GPT-5/framework | GPT-5-mini/simplified |
| Senior Clinical | SRCLIN | GPT-5/framework | GPT-5-mini/simplified |
| General | GEN | GPT-5/framework | GPT-5-mini/simplified |

The seven persona types were modeled on published CDA-AMC committee membership categories. CDA-AMC’s Procedures for Reimbursement Reviews specify that CDEC and pERC comprise clinicians (including disease-area specialists), health economists, patient representatives, ethicists, and drug plan managers. Our personas represent role-based analytical lenses (distinct reasoning perspectives that different expertise areas bring to evidence evaluation) rather than simulations of specific named individuals. A committee chair role was not modeled separately because the chair function in CDA-AMC committees is primarily procedural (managing discussion flow and building consensus) rather than representing a distinct analytical perspective.

#### Weighted Voting

Within each round, panelists cast votes that are weighted by prompt type:

- Structured prompt panelists (Instance 1): weight = 2.0
- Simplified panelists (Instance 2): weight = 1.0 Effective committee weight: 7 × 2.0 + 7 × 1.0 = 21

The weight differential reflects prompt sophistication rather than persona importance: Instance 1 panelists perform a structured multi-domain assessment following the CDA-AMC Deliberative Framework (clinical value, unmet need, economic evaluation, implementation), producing more comprehensive reasoning. Instance 2 panelists provide a complementary diversity signal through simplified role-focused prompts on a different model. All seven persona types are weighted equally.

The round winner is determined by weighted plurality voting:

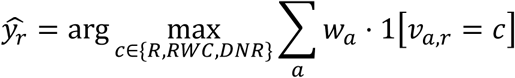

Tie-breaking follows a conservative ordering: DNR > RWC > R.

#### Probability Estimation

Let 𝐷_𝑟_ ∈ 𝑅, *RWC, DNR* be the committee outcome in round 𝑟𝑟, and 𝐷𝐷 the total rounds executed. The Monte Carlo estimate of the outcome distribution is:

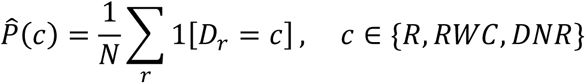

For binary evaluation (given the observed label space {RWC, DNR}):

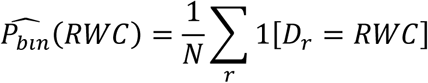

#### Convergence Criteria

The simulation runs until the probability estimate stabilizes or a maximum is reached:

**Table 4:**
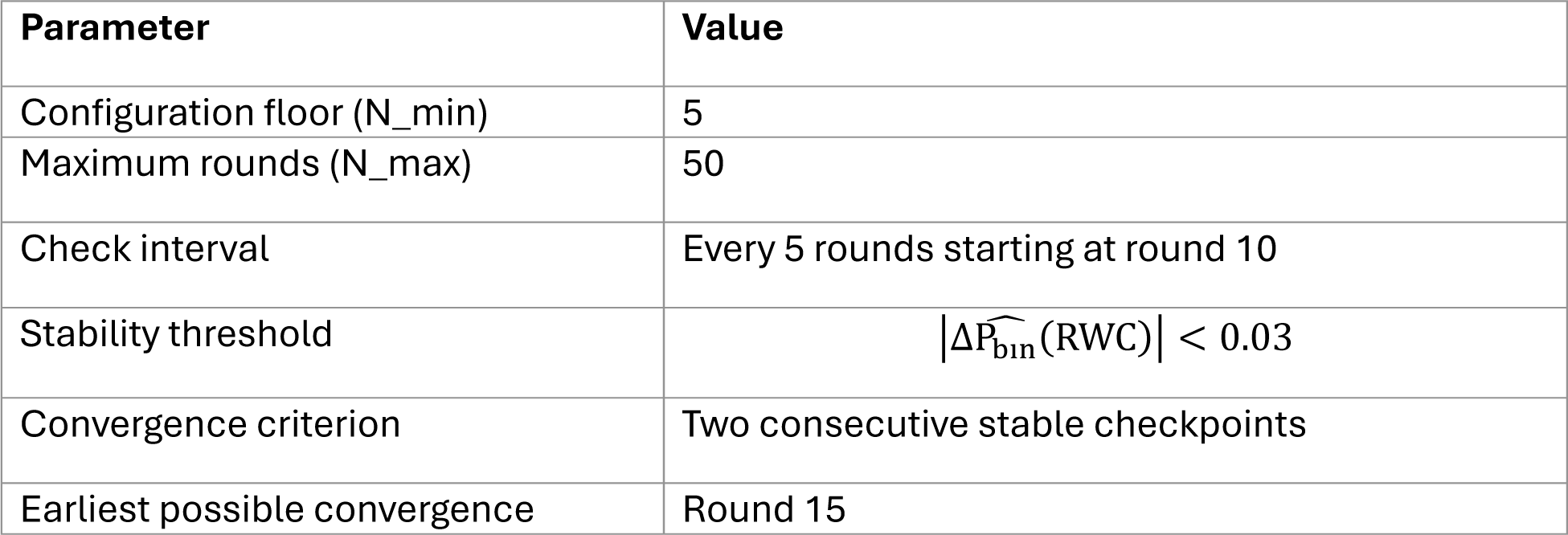
Convergence Specifics.

The configuration floor of 5 rounds ensures a minimum sample before any stability assessment. Convergence checks occur at 5-round intervals starting at round 10, requiring two consecutive stable checkpoints to terminate. Because the first checkpoint occurs at round 10 and the second at round 15, the earliest a simulation can converge is round 15.

#### Two-Axis Uncertainty Model

Prediction confidence requires both stability and consensus:

**Table 5:** Abstaining Specifics.

| Axis | Metrics | Question |
| --- | --- | --- |
| Stability (inter-round) | inter_round_margin,<br>inter_round_entropy | Does the winner flip across rounds? |
| Contestation (intra-round) | final_support,<br>vote_margin_median,<br>dissent_mean | Is each round's vote close? |

#### Uncertainty Metric Definitions

- final_support: Proportion of all weighted votes (across all rounds) cast for the predicted class
- vote_margin_median: Median across rounds of the within-round weighted vote margin between the top two classes
- inter_round_margin: Difference in proportion of rounds won by the top two classes
- dissent_mean: Mean proportion of panelist weight voting against the round winner, averaged across rounds

#### Selective Prediction (Abstention)

All abstention thresholds and Strength of Mandate cutoffs were prespecified on the calibration set (n=18) and frozen prior to test set evaluation. Thresholds were selected by identifying natural breakpoints in the distribution of uncertainty metrics that separated correct from incorrect predictions on the calibration set.

The system abstains when uncertainty exceeds pre-specified thresholds. Abstention occurs if ANY of:

- final_support < 0.60 (weak mandate)
- vote_margin_median < 0.14 (contested rounds)
- inter_round_margin < 0.40 (unstable winner)

**Strength of Mandate** classifies predictions into interpretable confidence levels:

**Table 6:** Confidence (mandate) Specifics.

| Category | Criteria | Interpretation |
| --- | --- | --- |
| High Mandate | `final_support` >= 0.80<br>AND<br>`vote_margin_median` >= 0.29 | Strong consensus |
| Contested | `final_support` in [0.60, 0.80) OR<br>`vote_margin_median` in [0.14, 0.29) | Stable but controversial |
| Weak Mandate | `final_support` < 0.60<br>OR<br>`vote_margin_median` < 0.14 | Low confidence |

This design enables a coverage-accuracy trade-off: by adjusting the ‘final_support’ threshold for abstention, users can increase prediction accuracy at the cost of reduced coverage (proportion of cases receiving a prediction). We report the coverage-accuracy curve to characterize this trade-off across operating points.

#### Conditions Prediction

For cases where the committee predicts RWC, each panelist’s structured vote includes boolean flags for 11 condition subcategories. Aggregation proceeds hierarchically:

1. **Panelist to Round**: Majority vote across 14 panelists for each subcategory
2. **Subcategory to Category**: Category = TRUE if ANY constituent subcategory is TRUE
3. **Round to Final**: Condition frequency = proportion of RWC rounds flagging each condition

Final prediction: Condition is predicted if frequency >= 0.50 across RWC rounds.

For condition text harmonization, we applied semantic clustering to group similar condition descriptions across panelists before frequency aggregation.

Conditions are evaluated only on cases with ground-truth recommendation = RWC (n=62), independent of the model’s predicted recommendation.

### Statistical Analysis

#### Hypothesis Testing

For the association between Strength of Mandate categories and prediction accuracy, we used a permutation test (10,000 iterations). The permutation test assesses whether the observed relationship between mandate category and correctness could have arisen by chance by repeatedly shuffling category labels and computing the chi-square statistic under the null distribution.

#### Evaluation Metrics

**Primary recommendation metrics:** - Accuracy - Macro-averaged F1 score (over observed labels {RWC, DNR}) - Sensitivity (RWC as positive class) - Specificity (DNR identification) - AUROC (using ̂*P_bin_* (*RWC*) as score)

**Calibration metrics:** - Expected Calibration Error (ECE) using 10 equal-width bins - Brier score using ̂*P_bin_* (*RWC*)

**Conditions metrics:** - Subset accuracy (exact match of all 5 categories) - Hamming accuracy - Per-category sensitivity, specificity, F1, PPV/NPV

#### Confidence Intervals

Bootstrap confidence intervals (1000 iterations) for all metrics.

#### Baselines

For recommendation prediction, we compared Monte Carlo Committee Simulation against a majority class baseline that always predicts Reimburse with Conditions. This baseline achieves maximum accuracy without discrimination (AUROC = 0.50) and establishes the class-imbalanced nature of the prediction task. Conditions prediction was evaluated on per-category metrics without baseline comparison due to severe class imbalance that renders aggregate accuracy comparisons uninformative.

#### Data and Code Availability

All data derived from publicly available CDA-AMC recommendation documents. Source PDFs are available from the CDA-AMC website.

## Results

### Study Population

Of 81 CDA-AMC recommendations published between October 2024 and December 2025, 67 met inclusion criteria after excluding 14 FMEC non-sponsored reviews (Table 1).

**Table 1.** Study Population Characteristics.

| Characteristic | Calibration Set | Test Set | Total |
| --- | --- | --- | --- |
| Sample size | 18 | 49 | 67 |
| Temporal range | Oct–Dec 2024 | Jan–Dec 2025 | Oct 2024–Dec 2025 |
| Committee |  |  |  |
| pERC (oncology) | 11 (61.1%) | 28 (57.1%) | 39 (58.2%) |
| CDEC (non-oncology) | 7 (38.9%) | 21 (42.9%) | 28 (41.8%) |
| Recommendation |  |  |  |
| Reimburse with Conditions | 17 (94.4%) | 45 (91.8%) | 62 (92.5%) |
| Do Not Reimburse | 1 (5.6%) | 4 (8.2%) | 5 (7.5%) |
| Reimburse | 0 (0%) | 0 (0%) | 0 (0%) |

Abstention thresholds were calibrated on recommendations from October–December 2024 (n=18), with primary evaluation conducted on recommendations from January–December 2025 (n=49). The class distribution reflects current HTA practice, where unconditional approval is rare and most novel therapies receive conditional reimbursement. This severe class imbalance (91.8% RWC in the test set) establishes a high baseline accuracy threshold that any predictive model must exceed to demonstrate genuine discriminative ability.

### Convergence

Simulations converged in a median of 15 rounds (range 15–40), with 85.7% of cases (42/49) reaching the stability criterion at the earliest possible checkpoint (round 15). Cases requiring more than 15 rounds to converge showed significantly lower accuracy than those converging early (57.1% vs 92.9%, Mann-Whitney U test, p = 0.006), suggesting prolonged deliberation reflects genuine prediction uncertainty rather than computational noise.

### Recommendation Prediction

Monte Carlo Committee Simulation achieved 93.2% accuracy (95% CI: 84.1–100.0%) on 44 submissions where the system expressed confidence in its prediction (Table 2). The system abstained from making a final decision on the remaining 5 submissions (10.2%) due to high uncertainty. Analysis of the 5 abstained cases showed 40.0% accuracy (2/5), confirming the system appropriately identified uncertain cases where its predictions were unreliable.

**Table 2.** Recommendation Prediction Performance.

| **Metric** | **Monte Carlo Committee Simulation** | **Majority Class Baseline** |
| --- | --- | --- |
| Accuracy, non-abstained, % (95% CI) | 93.2 (84.1–100.0) | — |
| Accuracy, all cases, % (95% CI) | 87.8 (79.6–95.9) | 91.8 (83.7–98.0) |
| Sensitivity for RWC, % (95% CI) | 91.1 (81.6–97.9) | 100.0 |
| Specificity for DNR, % (95% CI) | 50.0 (0.0–100.0) | 0.0 |
| AUROC (95% CI) | 0.817 (0.45–0.99) | 0.50 |
| ECE | 0.091 | — |
| Abstention rate, % | 10.2 | — |

*Abbreviations: RWC, Reimburse with Conditions; DNR, Do Not Reimburse; AUROC, area under the receiver operating characteristic curve; ECE, expected calibration error. Majority class baseline always predicts Reimburse with Conditions. 95% confidence intervals computed using 1000 bootstrap iterations (percentile method, seed=42).*

On the full test set including abstained cases, the system achieved 87.8% accuracy (95% CI: 79.6–95.9%), compared to 91.8% (95% CI: 83.7–98.0%) for the majority class baseline. The higher baseline accuracy reflects the extreme class imbalance; however, the committee approach demonstrated substantially better discrimination (AUROC 0.817 vs 0.50) and well-calibrated probability estimates (ECE = 0.091). Sensitivity for detecting Reimburse with Conditions was 91.1% (95% CI: 81.6–97.9%) and specificity for detecting Do Not Reimburse was 50.0% (95% CI: 0.0–100.0%). The system made 6 errors: 4 false negatives (RWC predicted as DNR) and 2 false positives (DNR predicted as RWC) (Table 3).

**Table 3.** Confusion Matrix (Test Set, n=49)

|  | **Predicted RWC** | **Predicted DNR** | **Total** |
| --- | --- | --- | --- |
| **Actual RWC** | 41 | 4 | 45 |
| **Actual DNR** | 2 | 2 | 4 |
| **Total** | 43 | 6 | 49 |

Prediction confidence, quantified by Strength of Mandate, was significantly associated with accuracy (permutation test, p = 0.0025). High mandate predictions achieved 96.8% accuracy (30/31), Contested predictions achieved 84.6% (11/13), and Weak mandate predictions achieved 40.0% (2/5) (Table 4). Notably, 5 of 6 errors (83.3%) occurred in cases with Contested or Weak mandate, confirming that the system’s uncertainty estimates identify difficult predictions. The small subgroup sizes (particularly n=5 for Weak Mandate) yield wide confidence intervals; while the monotonic trend across mandate categories is significant (p=0.0025), individual category estimates should be interpreted cautiously. By adjusting the abstention threshold, users can trade coverage for accuracy: at 70% coverage, accuracy reaches 97.1% (Table 5).

**Table 4.** Accuracy by Strength of Mandate.

| **Strength of Mandate** | **N (%)** | **Correct** | **Accuracy, %** |
| --- | --- | --- | --- |
| High | 31 (63.3%) | 30 | 96.8 |
| Contested | 13 (26.5%) | 11 | 84.6 |
| Weak | 5 (10.2%) | 2 | 40.0 |

*Permutation test (10,000 iterations), $p = 0.0025$ . Strength of Mandate classification based on final support rate, median vote margin, and mean dissent rate across Monte Carlo rounds.*

**Table 5.** Coverage-Accuracy Trade-off.

| **Coverage** | **N Predicted** | **Accuracy, %** | **Threshold (final support)** |
| --- | --- | --- | --- |
| 100% | 49 | 87.8 | 0.492 |
| 90% (default) | 44 | 93.2 | 0.603 |
| 80% | 39 | 94.9 | 0.676 |
| 70% | 34 | 97.1 | 0.705 |
| 60% | 29 | 96.6 | 0.835 |

*At the default 90% coverage threshold, abstaining on cases with lowest confidence improves accuracy from 87.8% to 93.2%, exceeding the majority class baseline.*

### Conditions Prediction

For recommendations correctly predicted as Reimburse with Conditions (n=41), the system predicted applicable condition categories with 48.8% subset accuracy. Subset accuracy requires the system to correctly predict all 5 condition categories simultaneously (Population Restrictions, Prescriber/Setting, Continuation Conditions, Economic Conditions, and Evidence Conditions) to count as correct. With 2^5^ = 32 possible category combinations, this is a stringent metric where a random baseline achieves only 3.1% exact match. The observed 48.8% provides actionable information for formulary negotiation preparation.

Performance varied by category (Table 6). Economic Conditions showed the highest accuracy (97.6%, F1 = 0.99), followed by Population Restrictions (90.2%, F1 = 0.95), Prescriber/Setting Requirements (75.6%, F1 = 0.85), and Continuation Conditions (68.3%, F1 = 0.77). Continuation Conditions demonstrated the strongest discriminative ability (AUROC 0.896, 95% CI: 0.79–0.98), reflecting sufficient class balance (35 positive, 6 negative) for reliable evaluation. Evidence Conditions (time-limited approval) were correctly identified as absent in all 41 cases, consistent with no time-limited recommendations in the evaluation set. Overall multi-label performance achieved Hamming accuracy of 86.3% compared to 25.8% for a no-conditions baseline.

**Table 6.**
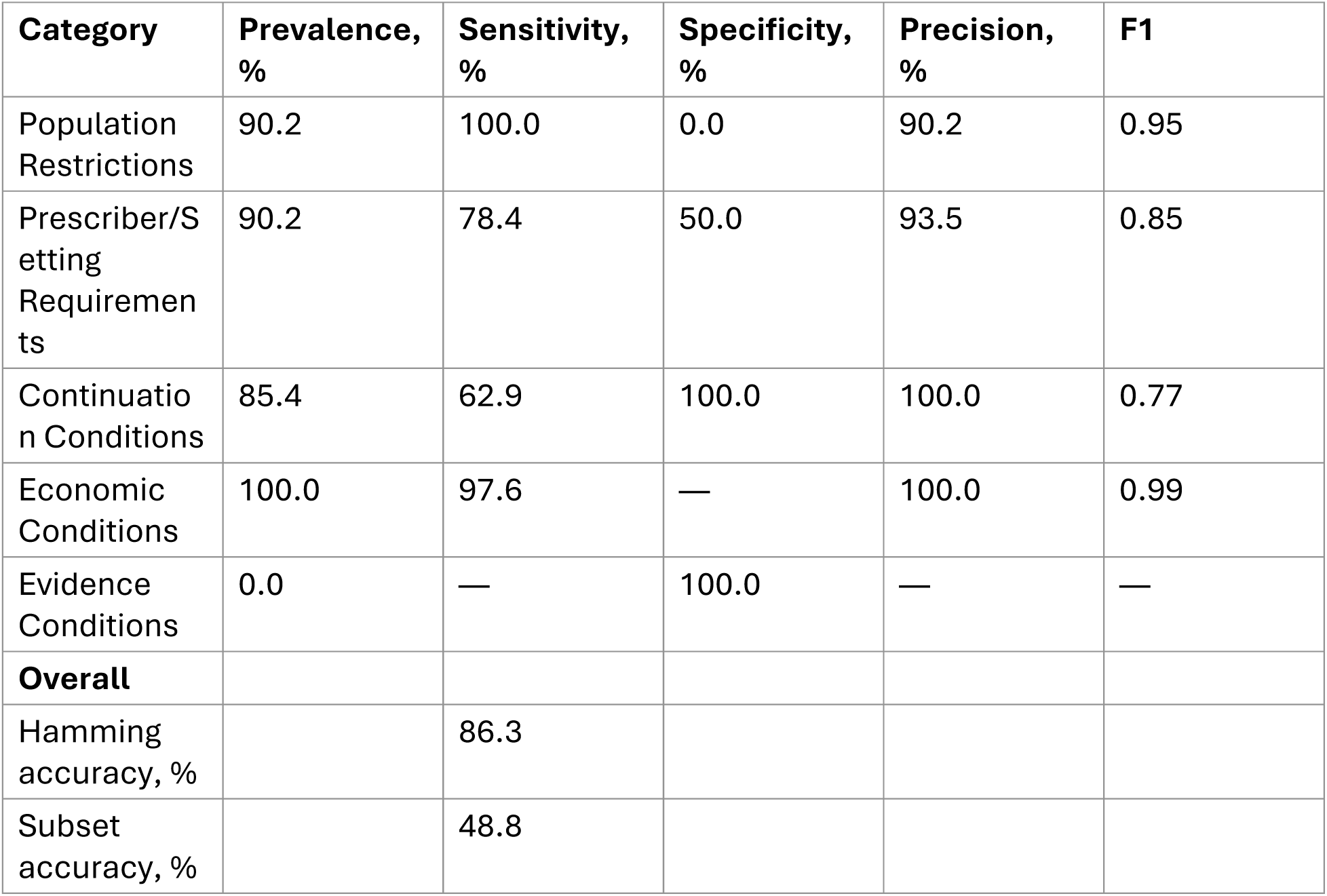
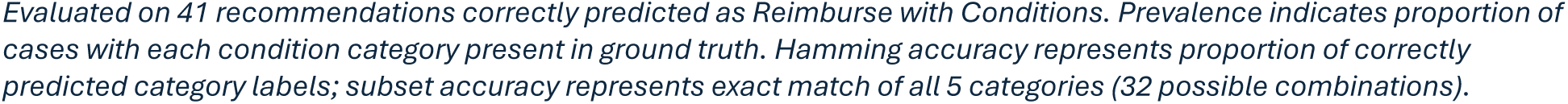
Conditions Prediction by Category.

## Discussion

### Principal Findings

We developed and evaluated Monte Carlo Committee Simulation, a neurosymbolic system for predicting Canadian drug reimbursement recommendations and their associated conditions, using a temporal external validation design where all predictions were made on recommendations issued after the knowledge cutoff of the underlying large language models. On submissions where the system expressed confidence, recommendation prediction achieved 93.2% accuracy, exceeding the majority class baseline. More importantly, the system demonstrated genuine discriminative ability (AUROC 0.817 vs 0.50) and well-calibrated uncertainty estimates that reliably identified difficult predictions. The Strength of Mandate metric stratified accuracy from 96.8% (High) to 40.0% (Weak), with 83.3% of errors occurring in cases the system flagged as uncertain. For condition prediction, the system achieved 48.8% subset accuracy—meaning it correctly predicted the exact combination of all 5 condition categories in nearly half of cases. This is a 32-class multi-label classification problem (2^5 possible combinations), where a random baseline achieves only 3.1% exact match. Correctly predicting whether a drug will face population restrictions AND prescriber requirements AND continuation criteria AND economic conditions AND evidence requirements, all simultaneously, provides sponsors with a specific actionable profile rather than a generic “conditional approval” label.

### Conditions Prediction as Novel Contribution

Prior work on HTA prediction spans two decades. Devlin and Parkin established that cost-effectiveness was the primary driver of NICE decisions;^19^; Dakin et al. extended this with multinomial models distinguishing “yes,” “no,” and “yes, but” outcomes, finding that cost-effectiveness alone correctly predicted 82% of decisions.^20^ Charokopou et al. analyzed 463 SMC submissions using logistic regression, identifying seven predictors including ICER thresholds and therapeutic area.^21^ More recent machine learning approaches include Wang et al.’s random forest models for SMC achieving >90% accuracy,^6^ Lin et al.’s multi-country analysis across five HTA agencies reaching 85-90% F1-scores,^22^, and Kanavos et al.’s analysis of 1,415 assessments across seven OECD countries, which notably found that 87.3% of decisions were positive with most involving restrictions.^23^ This class distribution aligns closely with our observed 92% RWC rate, suggesting our findings reflect broader HTA practice rather than CDA-AMC-specific patterns.

However, these traditional machine learning approaches share two limitations our work addresses. First, they require extensive manual feature extraction from submission documents, limiting scalability and introducing potential investigator bias in variable selection. Second, they predict only categorical outcomes, not the specific conditions attached to recommendations. Our LLM-based approach processes raw evidentiary documents without manual feature engineering, and the temporal external validation design addresses the data contamination concern unique to LLM applications (where historical HTA recommendations may be present in training corpora, making it impossible to distinguish reasoning from memorization).

To our knowledge, this represents the first attempt to prospectively predict the specific conditions attached to HTA reimbursement recommendations. Prior work on managed entry agreements and reimbursement conditions has been exclusively retrospective, developing taxonomies and analyzing patterns after decisions were made.^9,10^ The practical value of conditions prediction lies in enabling targeted preparation: knowing that a submission will likely face prior therapy requirements and price reduction conditions, rather than merely “conditional reimbursement,” allows sponsors to prepare negotiation strategies, anticipate formulary listing timelines, and allocate resources accordingly. The system’s ability to discriminate which submissions face which conditions, particularly for categories with sufficient class balance (Continuation Conditions AUROC 0.896), represents a meaningful advance over both categorical HTA prediction and prior taxonomic studies that characterized conditions only after decisions were made.

### Neurosymbolic Architecture

The Monte Carlo Committee Simulation architecture instantiates a neurosymbolic design where neural and symbolic components serve complementary functions. The neural components (14 persona-conditioned LLM panelists) handle evidence interpretation, drawing on the models’ encoded knowledge of clinical, economic, and policy domains to reason over unstructured evidentiary documents. The symbolic components (weighted voting, convergence criteria, abstention thresholds) perform collective inference with principled statistical properties, generating calibrated probability distributions and interpretable confidence metrics.

Neither component alone would suffice. Single-prompt LLM approaches cannot quantify uncertainty in a principled way; their outputs provide no indication of whether the model is confident or guessing. Conversely, purely symbolic approaches cannot process the lengthy, unstructured documents that characterize HTA submissions. The separation allows neural reasoning to handle ambiguity and implicit domain knowledge while symbolic aggregation provides transparency and calibration. Individual panelists achieved accuracies ranging from 40.8% to 91.8%, yet the ensemble substantially outperformed any individual, demonstrating that diversity in LLM instantiation, like diversity in human committees, enables robust collective judgment.

### Data Contamination and Temporal External Validation

A critical methodological challenge in validating LLM-based clinical and health economics applications is data contamination: the phenomenon where language models incorporate information from evaluation data in their training corpus, leading to inflated performance estimates.^24,25^ Many published benchmark and validation studies evaluate on data that may have been part of the model’s training corpus, creating apparent success that reflects memorization rather than genuine reasoning.^26–28^ This distinction matters profoundly for real-world deployment: when a system is used to inform decisions about truly novel submissions (the actual use case), performance may degrade substantially if prior success depended on pattern-matching to cached outcomes.

Consider the consequences of misplaced trust. A pharmaceutical sponsor might rely on an LLM-predicted “likely Reimburse with Conditions” for a novel therapy, allocating resources based on this forecast, only to discover the model was reproducing patterns from similar historical cases rather than reasoning about the submission’s specific evidence. An HTA agency might use such predictions for workload planning, unaware that accuracy estimates were inflated by contamination. Recent research has documented extensive benchmark leakage in LLM evaluations,^29,30^, with some studies finding that nearly all popular benchmarks show evidence of contamination.

Our temporal external validation design directly addresses this concern. By restricting evaluation to recommendations published after the LLMs’ knowledge cutoffs (GPT-5: September 30, 2024; GPT-5-mini: May 31, 2024), we ensure the system must reason from evidentiary documents rather than retrieve cached outcomes. The 93.2% accuracy on confident predictions and 0.817 AUROC were achieved on submissions the models could not have encountered during training.

This design comes at a cost. Using state-of-the-art models with recent knowledge cutoffs necessarily limits the available sample size for validation. Older models with longer post-cutoff evaluation periods are often deprecated by their providers. This creates a fundamental tension in LLM validation research that the field has not yet resolved: the models most likely to perform well are precisely those with the least available prospective data.

### Uncertainty Quantification as Decision Support

The system’s calibrated uncertainty enables a human-AI collaboration model rather than full automation. The Strength of Mandate classification stratified predictions into actionable confidence tiers: High Mandate cases (63.3% of submissions) achieved 96.8% accuracy and can be processed with minimal oversight; Contested cases (26.5%) achieved 84.6% accuracy and warrant careful review; Weak Mandate cases (10.2%) achieved only 40.0% accuracy and should trigger detailed manual review.

The coverage-accuracy trade-off empowers users to set risk tolerance explicitly. At the default 90% coverage, accuracy reaches 93.2%. Organizations requiring higher confidence can reduce coverage to 70%, achieving 97.1% accuracy while flagging 30% of submissions for human review. This flexibility accommodates diverse use cases, from rapid portfolio triage accepting some error to high-stakes individual submission assessment demanding near-certainty.

### Implications for Practice

**For pharmaceutical sponsors**, Monte Carlo Committee Simulation offers earlier signal on reimbursement trajectory during evidence development and submission preparation. The system’s condition predictions enable targeted preparation of negotiation strategies and reimbursement dossiers. Perhaps most valuably, the uncertainty quantification identifies submissions likely to face difficult committee deliberations, allowing sponsors to allocate expert resources where they are most needed.

**For HTA agencies and health systems**, the system offers decision support rather than decision replacement. Potential applications include consistency checking across recommendations, identifying cases that may warrant additional deliberation, and training new committee members on the factors that drive different outcomes. The transparent uncertainty quantification highlights which cases may prove contentious, allowing committees to anticipate where additional discussion or evidence review may be valuable.

### Strengths

This study has several methodological strengths. First, the temporal external validation design ensures that all predictions were made on recommendations the models could not have encountered during training, addressing the data contamination concern that undermines most LLM validation studies. Second, abstention thresholds were calibrated on a held-out calibration set and frozen before test evaluation, preventing threshold optimization on test data. Third, ground truth labels were obtained from official CDA-AMC structured data exports rather than manual annotation, eliminating labeling subjectivity for recommendation outcomes. Fourth, the two-level prediction framework (recommendations and conditions) provides more actionable information than binary classification alone. Fifth, the public availability of CDA-AMC source documents enables independent verification and replication.

### Limitations

Several limitations warrant consideration. First, the test set comprised only 49 recommendations, reflecting the real-world volume of HTA decisions but limiting statistical precision; confidence intervals are appropriately wide. Second, the severe class imbalance (91.8% RWC) means that aggregate accuracy comparisons are dominated by the majority class; we addressed this by reporting AUROC and by evaluating conditions without baseline comparison given category-level imbalances exceeding 85%. Third, evaluation was restricted to a single jurisdiction (CDA-AMC), though the condition taxonomy was designed for cross-jurisdictional generalizability. Fourth, with only 4 DNR cases in the test set, specificity estimates (50.0%, 95% CI: 0.0–100.0%) are highly uncertain. Fifth, detailed prompts are not disclosed due to commercial considerations, though they are available for academic replication upon request.

### Future Directions

Several directions merit further investigation. First, adapting the system to other HTA jurisdictions (NICE, PBAC, G-BA, INESSS) would require designing panelists that reflect each agency’s deliberative framework rather than simple external validation. Second, exploring open-source and locally-hosted language models would enable validation on larger samples with known, stable training cutoffs, addressing the fundamental tension between model capability and available prospective data. Third, prospective deployment studies with user feedback would assess the system’s value in actual decision support workflows. Fourth, extending the approach to other HTA decision types (medical devices, diagnostics, clinical pathways) would broaden applicability.

## Conclusion

Monte Carlo Committee Simulation with LLM panelists demonstrates that neurosymbolic architectures can achieve temporally validated prediction of HTA recommendations and their associated conditions with calibrated uncertainty. The system’s ability to identify when it might be wrong, and to abstain accordingly, positions it as a forecasting aid that complements human deliberation. This enables strategic preparation: anticipating specific condition types and allocating expert resources toward genuinely uncertain submissions. By ensuring evaluation on data the models could not have memorized, we provide evidence that large language models can genuinely reason about HTA evidence rather than merely reproduce historical patterns. Validation in additional jurisdictions and deployment studies assessing real-world utility represent important next steps toward integration into pharmaceutical development and health technology assessment workflows.

## Data Availability

https://www.cda-amc.ca/

## References

1. Bond K, Stiffell R, Ollendorf DA. Principles for deliberative processes in health technology assessment. International Journal of Technology Assessment in Health Care. 2020;36(4):445–452. doi:10.1017/S0266462320000550

2. Bombard Y, Abelson J, Simeonov D, Gauvin FP. Eliciting ethical and social values in health technology assessment: A participatory approach. Soc Sci Med. Jul 2011;73(1):135–44. doi:10.1016/j.socscimed.2011.04.017

3. Fleurence RL, Bian J, Wang X, et al. Generative Artificial Intelligence for Health Technology Assessment: Opportunities, Challenges, and Policy Considerations: An ISPOR Working Group Report. Value in Health. 2025/02/01/ 2025;28(2):175–183. 10.1016/j.jval.2024.10.3846

4. Zemplényi A, Tachkov K, Balkanyi L, et al. Recommendations to overcome barriers to the use of artificial intelligence-driven evidence in health technology assessment. Original Research. Frontiers in Public Health. 2023-April-26 2023;Volume 11 - 2023doi:10.3389/fpubh.2023.1088121

5. Ramezani M, Bakhtiari A, Daroudi R, et al. Applications of artificial intelligence and the challenges in health technology assessment: a scoping review and framework with a focus on economic dimensions. Health Economics Review. 2025/06/04 2025;15(1):46. doi:10.1186/s13561-025-00645-4

6. Wang Y, Tolley K, Francois C, Toumi M. Machine learning-based models for prediction of innovative medicine reimbursement decisions in Scotland. Journal of Epidemiology and Population Health. 2025/02/01/ 2025;73(1):202802. 10.1016/j.jeph.2024.202802

7. Maity S, Saikia MJ. Large Language Models in Healthcare and Medical Applications: A Review. Bioengineering (Basel). Jun 10 2025;12(6)doi:10.3390/bioengineering12060631

8. Artsi Y, Sorin V, Glicksberg BS, Korfiatis P, Nadkarni GN, Klang E. Large language models in real-world clinical workflows: a systematic review of applications and implementation. Front Digit Health. 2025;7:1659134. doi:10.3389/fdgth.2025.1659134

9. Garrison LP, Towse A, Briggs A, et al. Performance-Based Risk-Sharing Arrangements—Good Practices for Design, Implementation, and Evaluation: Report of the ISPOR Good Practices for Performance-Based Risk-Sharing Arrangements Task Force. Value in Health. 2013/07/01/ 2013;16(5):703-719. 10.1016/j.jval.2013.04.011

10. Walker S, Sculpher M, Claxton K, Palmer S. Coverage with evidence development, only in research, risk sharing, or patient access scheme? A framework for coverage decisions. Value Health. May 2012;15(3):570–9. doi:10.1016/j.jval.2011.12.013

11. Liu X, Chen T, Da L, Chen C, Lin Z, Wei H. Uncertainty Quantification and Confidence Calibration in Large Language Models: A Survey. presented at: Proceedings of the 31st ACM SIGKDD Conference on Knowledge Discovery and Data Mining V2; 2025; Toronto ON, Canada. 10.1145/3711896.3736569

12. Geng J, Cai F, Wang Y, Koeppl H, Nakov P, Gurevych I. A Survey of Confidence Estimation and Calibration in Large Language Models. Association for Computational Linguistics; 2024:6577–6595.

13. Kautz HA. The third AI summer: AAAI Robert S. Engelmore Memorial Lecture. AI Magazine. 2022;43(1):105–125. 10.1002/aaai.12036

14. Garcez AdA, Lamb LC. Neurosymbolic ai: The 3 rd wave. Artificial Intelligence Review. 2023;56(11):12387–12406.

15. Naderalvojoud B, Hernandez-Boussard T. Improving machine learning with ensemble learning on observational healthcare data. AMIA Annu Symp Proc. 2023;2023:521–529.

16. Wood D, Mu T, Webb AM, Reeve HWJ, Luján M, Brown G. A unified theory of diversity in ensemble learning. J Mach Learn Res. 2023;24(1):Article 359.

17. Cheng Y, Chang Y, Wu Y. A survey on data contamination for large language models. *arXiv preprint arXiv:250214425*. 2025;

18. Chen S, Chen Y, Li Z, et al. Benchmarking large language models under data contamination: A survey from static to dynamic evaluation. 2025:10091–10109.

19. Devlin N, Parkin D. Does NICE have a cost-effectiveness threshold and what other factors influence its decisions? A binary choice analysis. Health Econ. May 2004;13(5):437–52. doi:10.1002/hec.864

20. Dakin H, Devlin N, Feng Y, Rice N, O’Neill P, Parkin D. The Influence of Cost-Effectiveness and Other Factors on Nice Decisions. Health Econ. Oct 2015;24(10):1256–1271. doi:10.1002/hec.3086

21. Charokopou M, Majer IM, Raad J, Broekhuizen S, Postma M, Heeg B. Which factors enhance positive drug reimbursement recommendation in Scotland? A retrospective analysis 2006-2013. Value Health. Mar 2015;18(2):284–91. doi:10.1016/j.jval.2014.12.008

22. Lin R, Eaves K, Mills M, Kanavos P. HTA64 Leveraging machine learning for predicting health technology assessment outcomes. Value in Health. 2024;27(12, Supplement):S365–S365.

23. Kanavos P, Visintin E, Gentilini A. Algorithms and heuristics of health technology assessments: a retrospective analysis of factors associated with HTA outcomes for new drugs across seven OECD countries. Social Science & Medicine. 2023;331:116045.

24. Sainz O, Campos J, García-Ferrero I, Etxaniz J, de Lacalle OL, Agirre E. NLP evaluation in trouble: On the need to measure LLM data contamination for each benchmark. 2023:10776–10787.

25. Xu C, Guan S, Greene D, Kechadi M. Benchmark data contamination of large language models: A survey. *arXiv preprint arXiv:240604244*. 2024;

26. Sainz O, Campos J, García-Ferrero I, Etxaniz J, de Lacalle OL, Agirre E. NLP Evaluation in trouble: On the Need to Measure LLM Data Contamination for each Benchmark. Association for Computational Linguistics; 2023:10776–10787.

27. Deng C, Zhao Y, Tang X, Gerstein M, Cohan A. Investigating Data Contamination in Modern Benchmarks for Large Language Models. Association for Computational Linguistics; 2024:8706–8719.

28. Balloccu S, Schmidtová P, Lango M, Dusek O. Leak, Cheat, Repeat: Data Contamination and Evaluation Malpractices in Closed-Source LLMs. Association for Computational Linguistics; 2024:67–93.

29. Deng C, Zhao Y, Tang X, Gerstein M, Cohan A. Investigating data contamination in modern benchmarks for large language models. 2024:8706–8719.

30. Chen S, Chen Y, Li Z, et al. Recent advances in large langauge model benchmarks against data contamination: From static to dynamic evaluation. *arXiv preprint arXiv:250217521*. 2025;

